# Association between SSRI Antidepressant Use and Reduced Risk of Intubation or Death in Hospitalized Patients with Coronavirus Disease 2019: a Multicenter Retrospective Observational Study

**DOI:** 10.1101/2020.07.09.20143339

**Authors:** Nicolas Hoertel, Marina Sánchez Rico, Raphaël Vernet, Nathanaël Beeker, Anne-Sophie Jannot, Antoine Neuraz, Elisa Salamanca, Nicolas Paris, Christel Daniel, Alexandre Gramfort, Guillaume Lemaitre, Mélodie Bernaux, Ali Bellamine, Cédric Lemogne, Guillaume Airagnes, Anita Burgun, Frédéric Limosin, On behalf of AP-HP / Universities / INSERM Covid-19 research collaboration and AP-HP Covid CDR Initiative

## Abstract

**Objective:** To examine the association between antidepressant use and the risk of intubation or death in hospitalized patients with COVID-19.

**Design:** Multicenter observational retrospective cohort study.

**Setting:** Greater Paris University hospitals, France.

**Participants:** 7,345 adults hospitalized with COVID-19 between 24 January and 1 April 2020, including 460 patients (6.3%) who received an antidepressant during the visit.

**Data source:** Assistance Publique-Hôpitaux de Paris Health Data Warehouse.

**Main outcome measures:** The primary endpoint was a composite of intubation or death. We compared this endpoint between patients who received antidepressants and those who did not in time-to-event analyses adjusting for patient characteristics (such as age, sex, and comorbidities), disease severity and other psychotropic medications. The primary analyses were multivariable Cox models with inverse probability weighting.

**Results:** Over a mean follow-up of 18.5 days (SD=27.1), 1,331 patients (18.1%) had a primary end-point event. Unadjusted hazard ratio estimates of the association between antidepressant use and the primary outcome stratified by age (i.e., 18-50, 51-70, 71-80, and 81+) were non-significant (all p>0.072), except in the group of patients aged 71-80 years (HR, 0.66; 95% CI, 0.45 to 0.98; p=0.041). Following adjustments, the primary analyses showed a significant association between use of any antidepressant (HR, 0.64; 95% CI, 0.51 to 0.80; p<0.001), SSRI (HR, 0.56; 95% CI, 0.42 to 0.75; p<0.001), and SNRI (HR, 0.57; 95% CI, 0.34 to 0.96; p=0.034), and reduced risk of intubation or death. Specifically, exposures to escitalopram, fluoxetine, and venlafaxine were significantly associated with lower risk of intubation or death (all p<0.05). These associations remain significant in multiple sensitivity analyses, except for the association between SNRI use and the outcome.

**Conclusions:** SSRI use could be associated with lower risk of death or intubation in hospitalized patients with COVID-19. Double-blind controlled randomized clinical trials of these medications for COVID-19 are needed.

**What is already known on this topic:**

- A prior meta-analysis, mainly including studies on selective serotonin reuptake inhibitors (SSRIs), showed that antidepressant use in major depressive disorder was associated with reduced levels of several pro-inflammatory cytokines, including IL-6, TNF-*α*, and CCL-2, which have been suggested to be associated with severe COVID-19.
- A recent in-vitro study supports antiviral effects of the SSRI fluoxetine on SARS-CoV-2.
- To our knowledge, no study has examined the efficacy of antidepressants in patients with COVID-19.

**What this study adds:**

- In a multicenter observational retrospective study, we examined the association between antidepressant use and the risk of intubation or death in hospitalized patients with COVID-19, adjusting for patient characteristics, disease severity and other psychotropic medications.
- Antidepressant use was significantly and substantially associated with reduced risk of intubation or death.
- At the level of antidepressant classes, SSRI use was significantly and substantially associated with reduced risk of intubation or death, but not other antidepressant classes.
- At the level of antidepressant medications, exposures to the SSRIs fluoxetine and escitalopram, and the SNRI venlafaxine were significantly associated with lower risk of intubation or death.
- Double-blind controlled randomized clinical trials of these medications for COVID-19 are needed.

## 1. Introduction

Global spread of the novel coronavirus SARS-CoV-2, the causative agent of coronavirus disease 2019 (COVID-19), has created an unprecedented infectious disease crisis worldwide. In the current absence of a vaccine or curative treatment with published evidence-based clinical efficacy, the search for an effective treatment for patients with COVID-19 among all available medications is urgently needed.^1,2^

Both COVID-19 and severe acute respiratory syndrome (SARS) are characterized by an overexuberant inflammatory response^3^ and, for COVID-19, viral load is associated with the worsening of symptoms.^4^

Prior work suggests a significant reduction of overactive inflammatory processes observed in individuals with major depressive disorder following antidepressant treatment.^5 6^ A recent meta-analysis^5^ of studies conducted in this population, mainly including selective serotonin reuptake inhibitors (SSRIs), supports that antidepressant use is associated with reduced plasma levels of several pro-inflammatory cytokines, including IL-6, TNF-*α*, and CCL-2, which have been suggested to be associated with severe COVID-19. These anti-inflammatory effects could be stronger for SSRIs, which could more potently inhibit microglial TNF-α and NO production through cAMP signaling regulation, than serotonin–norepinephrine reuptake inhibitors (SNRIs).^8^

A recent in-vitro study^9^ also supports antiviral effects of fluoxetine on SARS-CoV-2, although this effect was not observed for other SSRIs, including paroxetine and escitalopram.

In this context, we hypothesized that antidepressants, and more specifically SSRIs, could be potentially effective in reducing the risk of intubation or death in patients with COVID-19. Short-term use of low to moderate doses of antidepressants, and particularly of SSRIs, is generally well tolerated,^10^ and notably in older adults,^11 12^ who are the most prone to developing severe COVID-19.^13^

To our knowledge, no study has examined to date the efficacy of antidepressants for COVID-19. Observational studies of patients with COVID-19 taking medications for other indications can help determine their efficacy for COVID-19, decide which should be prioritized for randomized clinical trials, and minimize the risk for patients of being exposed to potentially harmful and ineffective treatments.

We took advantage of the Assistance Publique-Hôpitaux de Paris (AP-HP) Health Data Warehouse, which includes data on all patients with COVID-19 who had been consecutively admitted to any of the 39 Greater Paris University hospitals.

In this report, we examined the association between antidepressant use and the risk of intubation or death among adult patients who have been admitted to these medical centers with COVID-19. If a significant protective association were found, we sought to examine the associations between the use of different classes of antidepressants (i.e. SSRIs, SNRIs, tricyclic, tetracyclic and *α*2-antagonist antidepressants) and individual antidepressant medications, with this risk. We hypothesized that antidepressant use, and specifically SSRI use, would be associated with reduced risk of intubation or death in time-to-event analyses adjusting for potential confounders, including sex, age, obesity, current smoking status, any medical condition associated with increased risk of severe COVID-19, any medication prescribed according to compassionate use or as part of a clinical trial, markers of clinical and biological severity of COVID-19 at hospital admission, the presence of current mood or anxiety disorder or other psychiatric disorder, and use of any benzodiazepine or Z-drug, mood stabilizer, and antipsychotic medication.

## 2. Methods

### 2.1. Setting

We conducted this study at AP-HP, which comprises 39 hospitals, 23 of which are acute, 20 adult and 3 pediatric hospitals. We included all adults aged 18 years or over who have been admitted with COVID-19 to these medical centers from the beginning of the epidemic in France, i.e. January 24^th^, until April 1^st^. COVID-19 was ascertained by a positive reverse-transcriptase-polymerase-chain-reaction (RT-PCR) test from analysis of nasopharyngeal or oropharyngeal swab specimens. This observational study using routinely collected data received approval from the Institutional Review Board of the AP-HP clinical data warehouse (decision CSE-20-20_COVID19, IRB00011591). AP-HP clinical Data Warehouse initiative ensures patients’ information and informed consent regarding the different approved studies through a transparency portal in accordance with European Regulation on data protection and authorization n°1980120 from National Commission for Information Technology and Civil Liberties (CNIL). All procedures related to this work adhered to the ethical standards of the relevant national and institutional committees on human experimentation and with the Helsinki Declaration of 1975, as revised in 2008.

### 2.2. Data sources

We used data from the AP-HP Health Data Warehouse (‘Entrepôt de Données de Santé (EDS)’). This anonymized warehouse contains all the clinical data available on all inpatient visits for COVID-19 to any of the 39 Greater Paris University hospitals. The data obtained included patients’ demographic characteristics, vital signs, laboratory test and RT-PCR test results, medication administration data, past and current medication lists, past and current diagnoses, discharge disposition, ventilator use data, and death certificates.

### 2.3. Variables assessed

We obtained the following data for each patient at the time of the hospitalization: sex; age, which was categorized based on the OpenSAFELY study results^13^ (i.e. 18-50, 51-70, 71-80, 81+); obesity, defined as having a body-mass index higher than 30 kg/m^2^ or an International Statistical Classification of Diseases and Related Health Problems (ICD-10) diagnosis code for obesity (E66.0, E66.1, E66.2, E66.8, E66.9); self-reported current smoking status; any medical condition associated with increased risk of severe COVID-19^13-16^ based on ICD-10 diagnosis codes, including diabetes mellitus (E11), diseases of the circulatory system (I00-I99), diseases of the respiratory system (J00-J99), neoplasms (C00-D49), and diseases of the blood and blood-forming organs and certain disorders involving the immune mechanism (D5-D8); any medication prescribed according to compassionate use or as part of a clinical trial (e.g. hydroxychloroquine, azithromycin, remdesivir, tocilizumab, dexamethasone, or sarilumab); clinical severity of COVID-19 at admission, defined as having at least one of the following criteria^17^: respiratory rate > 24 breaths/min or < 12 breaths/min, resting peripheral capillary oxygen saturation in ambient air < 90%, temperature > 40°C, or systolic blood pressure <100 mm Hg; and biological severity of COVID-19 at admission, defined as having at least one of the following criteria:^17^,^18^ high neutrophil-to-lymphocyte ratio or low lymphocyte-to-C-reactive protein ratio (both variables were dichotomized at the median of the values observed in the full sample), or plasma lactate levels higher than 2 mmol/L. To take into account possible confounding by indication bias for antidepressants, we recorded whether patients had any current mood or anxiety disorder (F30- F48) or any other current psychiatric disorder (F00-F29 and F50-F99) based on ICD-10 diagnosis codes, and whether they were prescribed any benzodiazepine or Z-drug, any mood stabilizer (i.e. lithium or antiepileptic medications with mood stabilizing effects), or any antipsychotic medication.

All medical notes and prescriptions are computerized in Greater Paris University hospitals. Medications and their mode of administration (i.e., dosage, frequency, date, condition of intake) were identified from medication administration data or scanned handwritten medical prescriptions, through two deep learning models based on BERT contextual embeddings,^19^ one for the medications and another for their mode of administration. The model was trained on the APmed corpus,^20^ a previously annotated dataset for this task. Extracted medications names were then normalized to the Anatomical Therapeutic Chemical (ATC) terminology using approximate string matching.

### 2.4. Antidepressant use

Study baseline was defined as the date of hospital admission. Antidepressant use was defined as receiving any antidepressant at any time during the follow-up period, from study baseline to the end of the hospitalization or intubation or death.

### 2.5. Endpoint

The primary endpoint was the time from study baseline to intubation or death. For patients who died after intubation, the timing of the primary endpoint was defined as the time of intubation. Patients without an end-point event had their data censored on May 20^th^, 2020. Secondary endpoints included times from study baseline to intubation and death, separately.

### 2.6. Statistical analysis

We calculated frequencies and means (± standard deviations (SD)) of each baseline characteristic described above in patients receiving or not receiving antidepressants and compared them using chi-square tests or Welch’s t-tests.

To examine the associations between the use of any antidepressant, each class of antidepressants, and individual medications with the composite endpoint of intubation or death, we performed Cox proportional-hazards regression models. To help account for the nonrandomized prescription of antidepressants and reduce the effects of confounding, the primary analyses used propensity score analysis with inverse probability weighting.^21^ ^22^ The individual propensities for exposure were estimated by multivariable logistic regression models that included sex, age, obesity, smoking status, any medical condition associated with increased risk of severe COVID-19, any medication prescribed according to compassionate use or as part of a clinical trial, markers of clinical and biological severity of COVID-19 at hospital admission, the presence of mood or anxiety or other current psychiatric disorder, and any prescribed benzodiazepine or Z-drug, mood stabilizer, and antipsychotic medication. In the inverse-probability-weighted analyses, the predicted probabilities from the propensity-score models were used to calculate the stabilized inverse-probability-weighting weights.^21^ Associations between any antidepressant, each class of antidepressants, and individual treatments with the primary endpoint were then estimated using multivariable Cox regression models including the inverse-probability-weighting weights. Kaplan-Meier curves were performed using the inverse-probability-weighting weights,^23^ and their pointwise 95% confidence intervals were estimated using the nonparametric bootstrap method.^24^

We conducted sensitivity analyses, including multivariable Cox regression models comprising as covariates the same variables as the inverse-probability-weighted analyses, and univariate Cox regression models in matched analytic samples. For this latter analysis, we decided *a priori* to select one control for each exposed case for exposures to any antidepressant and each class of antidepressants, and two controls for each exposed case for individual antidepressant medications, based on the same variables used for both the inverse-probability-weighted and the multivariable Cox regression analyses. Weighted Cox regression models were used when proportional hazards assumption was not met. To reduce the effects of confounding, optimal matching was used in order to obtain the smallest average absolute distance across all clinical characteristics between exposed patient and non-exposed matched controls. We also performed multivariable Cox regression models including interaction terms to examine whether the association between antidepressant use and the primary endpoint significantly differed across subgroups defined by baseline characteristics.

Within patients exposed to antidepressants, we tested the association of daily dose (converted into fluoxetine-equivalent dose^25^ and dichotomized at the median value) with the primary endpoint. Furthermore, we examined whether exposure to a combination of antidepressants was associated with a significantly different risk of intubation or death than exposure to only one antidepressant.

We reproduced these analyses (i) among patients with critical COVID-19 hospitalized in intensive care units (ICUs), and (ii) using death and intubation as separate endpoints. Finally, among patients with COVID-19 not using antidepressants during the hospitalization, we examined whether the risk of intubation or death differed between patients receiving or not antidepressants in the 3 months before hospital admission.

For all associations, we performed residual analyses to assess the fit of the data, check assumptions, including proportional hazards assumptions, and examined the potential influence of outliers. To improve the quality of result reporting, we followed the recommendations of The Strengthening the Reporting of Observational Studies in Epidemiology (STROBE) Initiative.^26^ Our main analysis was focused on the association between antidepressant use and the composite outcome of intubation or death. We planned to examine the association between

Because analyses were exploratory, statistical significance was fixed *a priori* at two-sided p-value<0.05. Only if a significant protective association were found, we planned to examine the associations between the use of different classes of antidepressants (i.e. SSRIs, SNRIs, tricyclic, tetracyclic and α2-antagonist antidepressants) and individual antidepressant medications, with this risk. All analyses were conducted in R software version 2.4.3 (R Project for Statistical Computing).

## 3. Results

### 3.1. Characteristics of the cohort

Of the 9,509 patients with a positive COVID-19 RT-PCR test consecutively admitted to the hospital, a total of 2,164 patients (22.8%) were excluded because of missing data or their young age (i.e. less than 18 years old of age). Of the remaining 7,345 adult inpatients, 460 patients (6.3%) received an antidepressant during the hospitalization, at a mean fluoxetine-equivalent dose of 21.4 mg (SD=13.6) per day (**Figure 1**). Doses of each antidepressant are shown in **eTable 1**. Among patients exposed to antidepressants, 391 (85.0%) were exposed to only one antidepressant, 69 (14.1%) received two antidepressants, and 4 (0.9%) patients were exposed to more than two different antidepressants. Mean age of patients exposed to antidepressants was 74.8 (SD=15.5) years, whereas it was 56.8 (SD=19.3) years in those who were not (Welch’s t-test=-23.70, p<0.001).

**Figure 1.**
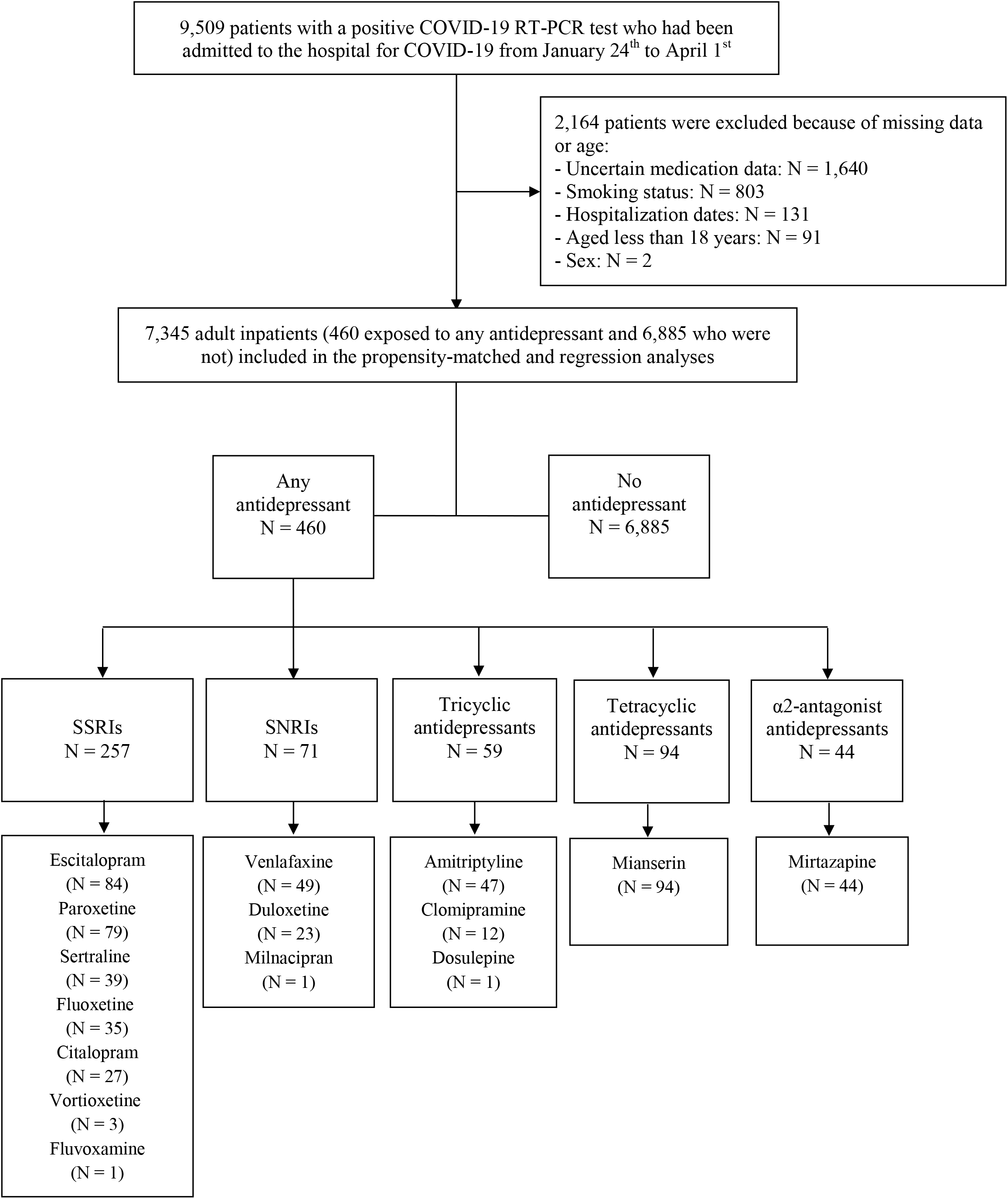
Study cohort. Antidepressant exposure was defined as receiving an antidepressant at any time during the follow-up period, from hospital admission until the end of the hospitalization or intubation or death. All intubated patients exposed to an antidepressant received it before the intubation in the study. SSRIs, selective serotonin reuptake inhibitors; SNRIs, serotonin-norepinephrine reuptake inhibitors.

COVID-19 RT-PCR test results were obtained after a median delay of 1 day (SD=12.1) from the date of hospital admission. This delay was not significantly different between patients receiving or not receiving antidepressants [median in the exposed group=1 day (SD=14.1); median in the non-exposed group=1 day (SD=11.9); Mood’s median test Chi-square=0.21, p=0.650)].

Over a mean follow-up of 18.5 days (SD=27.1; median=4 days; range: 1 day to 117 days), 1,331 patients (18.1%) had a primary end-point event at the time of data cutoff on May 20^th^. In patients exposed to antidepressants, the mean follow-up was 20.1 days (SD=23.2; median=11 days; range: 1 day to 112 days), while it was of 18.4 days (SD=27.4; median=4 days; range: 1 day to 117 days) in those who were not. The times to follow-up by treatment exposure are shown in **eTable 2**.

All baseline characteristics were independently and significantly associated with the primary endpoint, except for smoking, any medication prescribed according to compassionate use or as part of a clinical trial, any current mood or anxiety disorder, and any other current psychiatric disorder (**Table 1**).

**Table 1.**
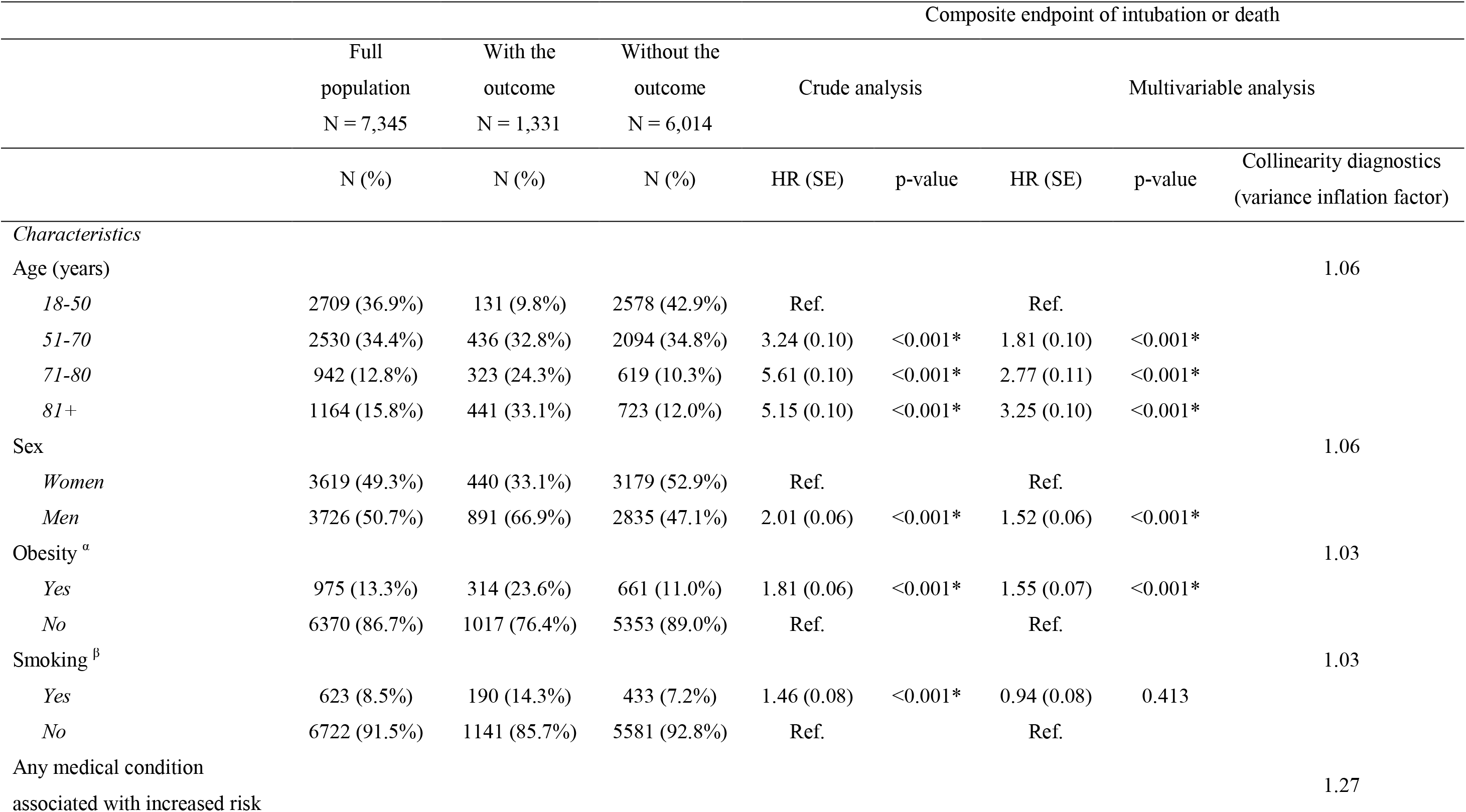
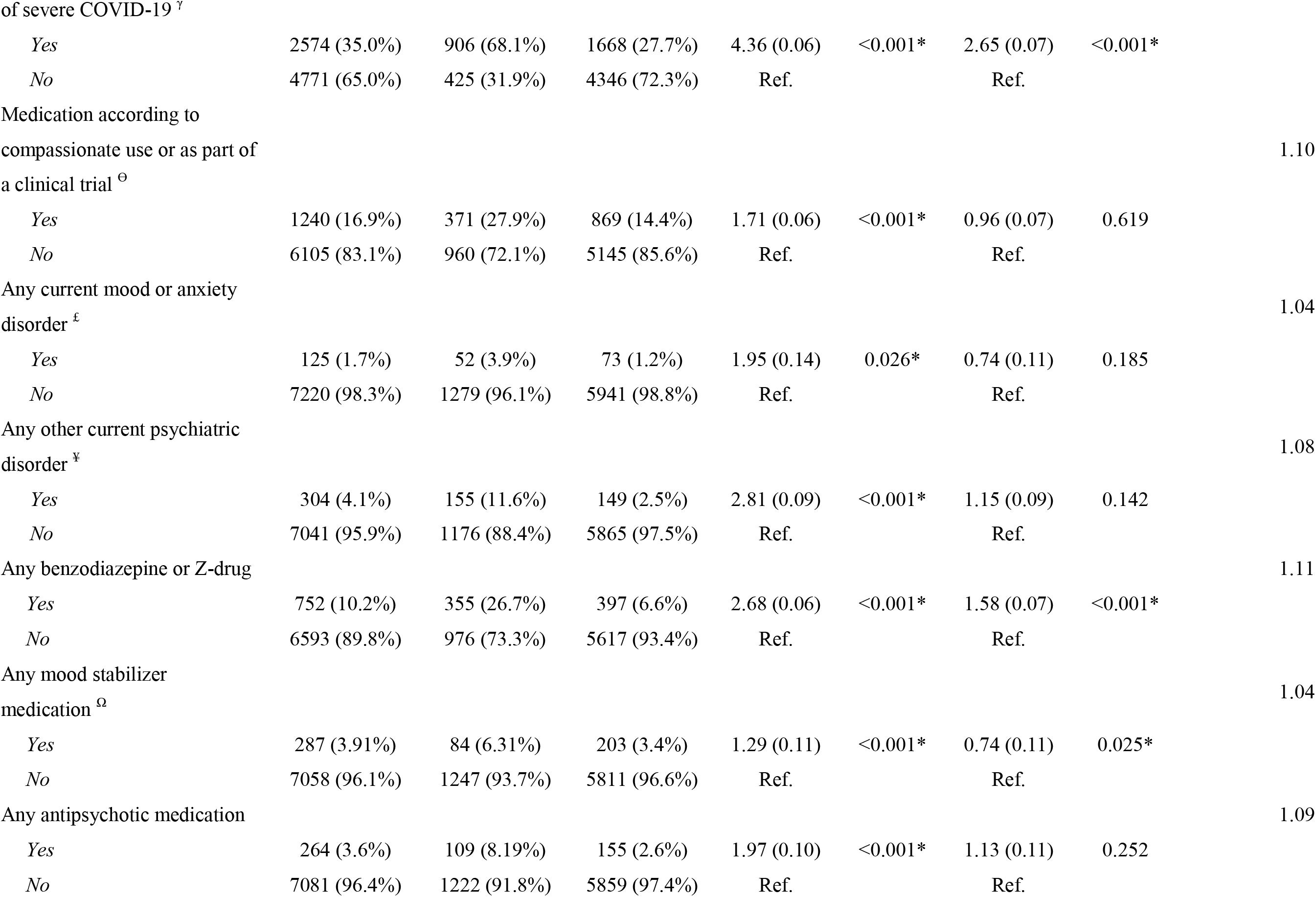
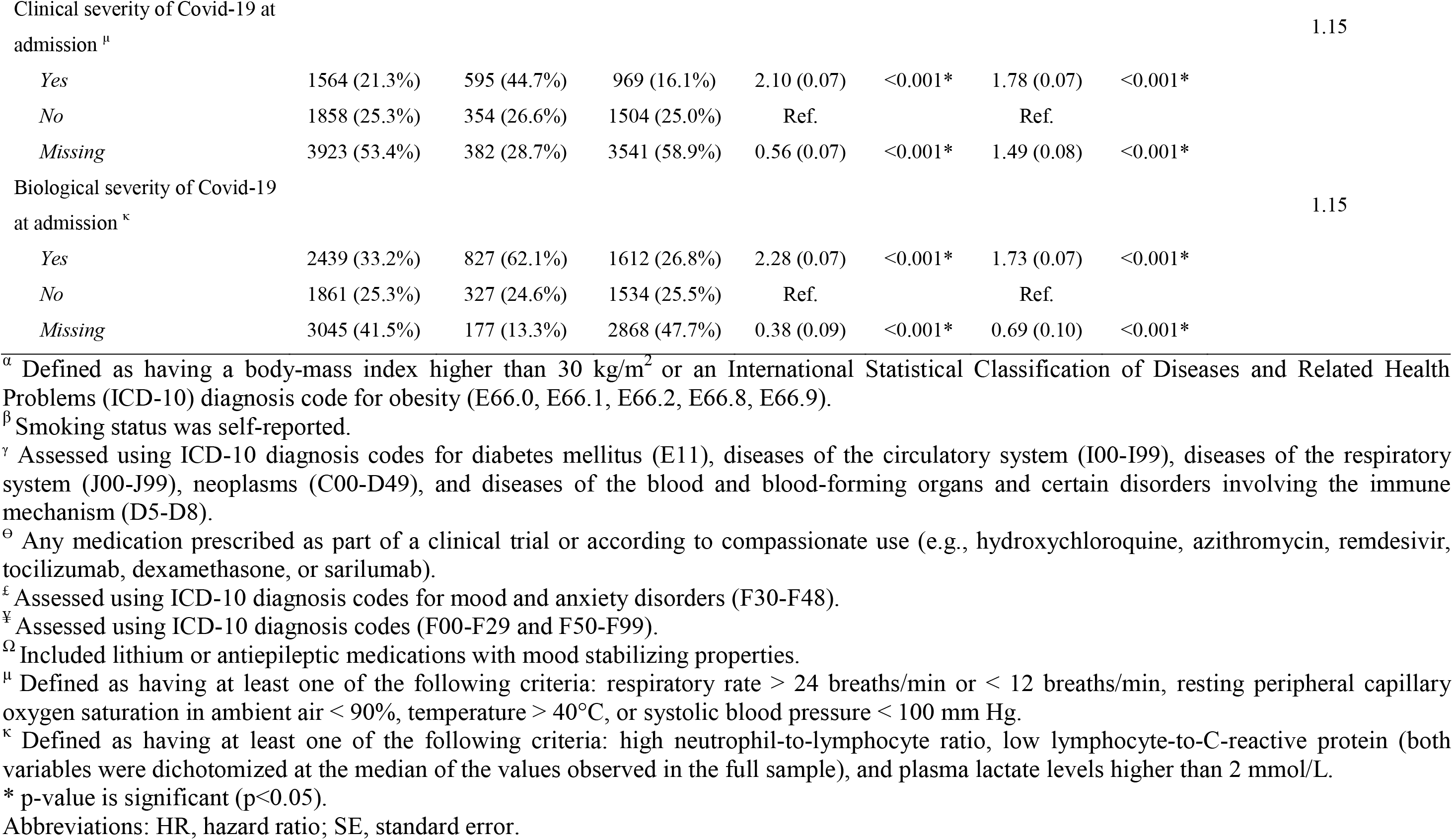
Associations of baseline clinical characteristics with the composite endpoint of intubation or death in the cohort of adult patients who had been admitted to the hospital with Covid-19 (N=7,345).

The distributions of the patients’ characteristics according to antidepressant exposure, and exposure by class of antidepressants and by individual medication are shown in **eTables 3 to 13**. In the full sample, antidepressant use significantly differed according to all baseline characteristics, as did use of each antidepressant class and individual medication. The direction of these associations indicated older age and overall greater medical severity of patients receiving antidepressants and each antidepressant class than those who did not receive any antidepressant. After applying the propensity score weights, these differences were substantially reduced (**eTables 3 to 13**). In the matched analytic samples, there were no significant differences in patients’ characteristics across the different exposures, except for antidepressant use, for which sex ratio significantly differed (**eTables 3 to 13**).

### 3.2. Study endpoint

Among patients receiving any antidepressant, SSRIs, SNRIs, tricyclic antidepressants, tetracyclic antidepressants, and α2-antagonist antidepressants, the primary endpoint of intubation or death occurred respectively in 143 patients (31.1%), 75 patients (29.2%), 18 patients (25.4%), 19 patients (32.2%), 43 patients (45.7%) and 12 patients (27.3%), while 1,188 non-exposed patients (17.3%) had this outcome (**Table 2**). Unadjusted hazard ratio estimates of the association between antidepressant use and the primary outcome stratified by age (i.e., 18-50, 51-70, 71-80, and 81+) were non-significant (all p>0.072), except in the group of patients aged 71-80 years where antidepressant use was significantly associated with lower risk of intubation or death (HR, 0.66; SE, 0.20; p=0.041) (**eTable 20**). When adjusting for the older age and the greater medical severity of patients who received an antidepressant than those who did not, the primary multivariable analyses with inverse probability weighting showed a significant association between use of any antidepressant (HR, 0.64; 95% CI, 0.51 to 0.80, p<0.001) (**Figure 2**), any SSRI (HR, 0.56; 95% CI, 0.42 to 0.75, p<0.001), and any SNRI (HR, 0.57; 95% CI, 0.34 to 0.96, p=0.034) (**Figure 3**), and reduced risk of intubation or death. There were no significant associations between this outcome and exposure to tricyclic antidepressants (HR, 0.96; 95% CI, 0.57 to 1.61, p=0.880), tetracyclic antidepressants (HR, 0.87; 95% CI, 0.61 to 1.25, p=0.454), and α2-antagonist antidepressants (HR, 0.29; 95% CI, 0.28 to 1.02, p=0.058) (**Table 2; Figure 3**). Specifically, exposure to escitalopram (HR, 0.61; 95% CI, 0.40 to 0.92, p=0.018), fluoxetine (HR, 0.32; 95% CI, 0.14 to 0.73, p=0.007), or venlafaxine (HR, 0.47; 95% CI, 0.24 to 0.91, p=0.025) was significantly associated with a reduced risk of intubation or death. There were no significant differences in this risk according to exposure to any other molecule (**Table 2; eFigures 1 to 5**). The association between antidepressant exposure and the outcome did not significantly differ across subgroups defined by baseline characteristics, except for markers of biological severity of COVID-19 at hospital admission (**eTable 14**).

**Figure 2.**
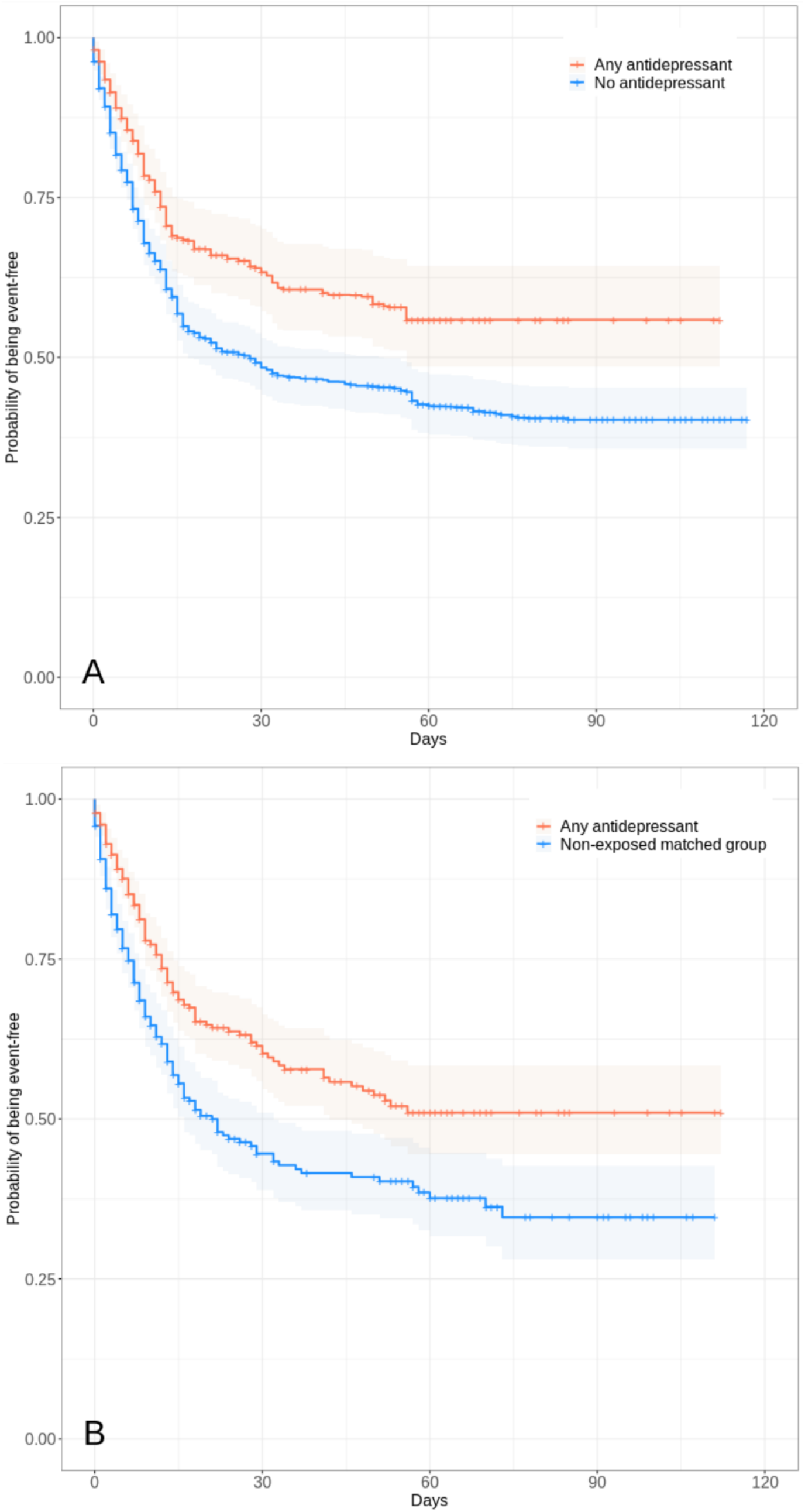
Kaplan-Meier curves of the composite endpoint of intubation or death in the full sample (N=7,345) (A) and in the matched analytic sample (N=920) (B) of patients who had been admitted to the hospital with Covid-19, according to antidepressant exposure. The shaded areas represent pointwise 95% confidence intervals.

**Figure 3.**
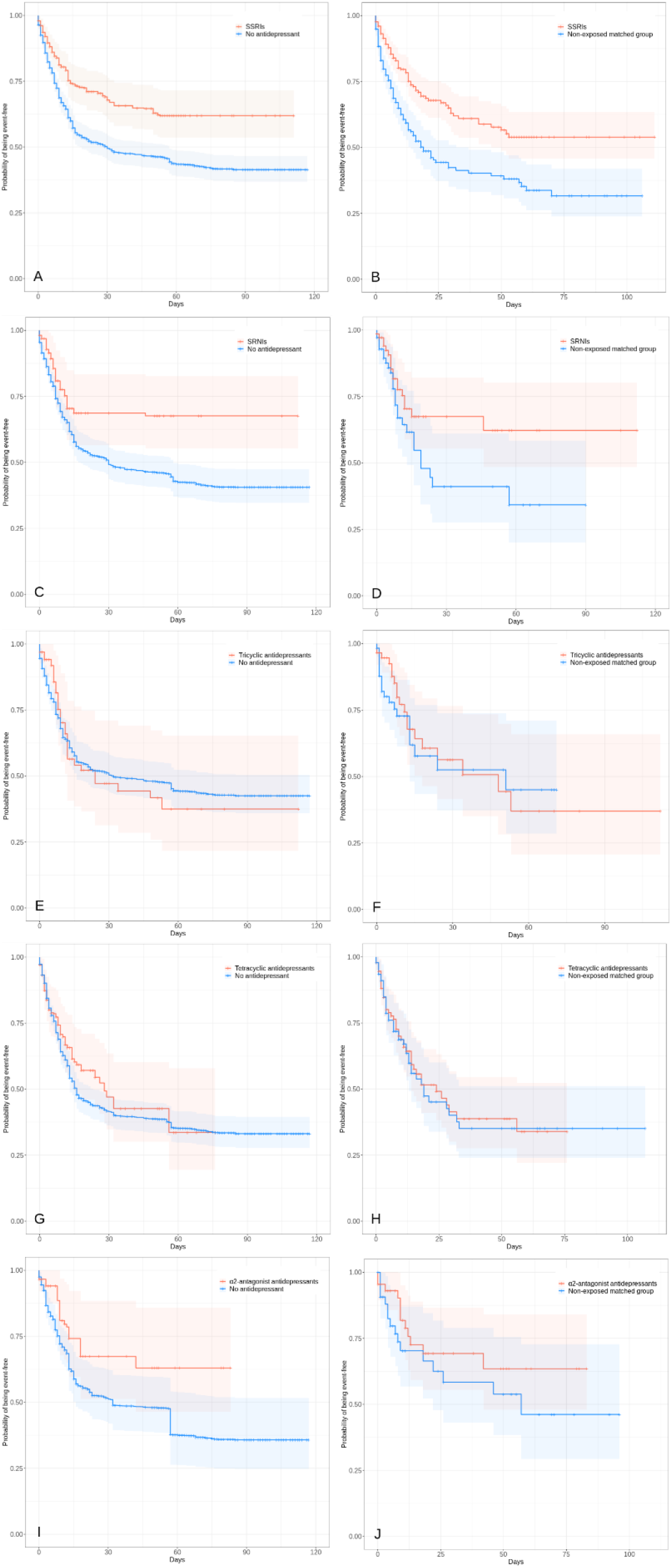
Kaplan-Meier curves of the composite endpoint of intubation or death in the full sample (A, C, E, G, and I) and in the matched analytic samples (B, D, F, H, and J) of patients who had been admitted to the hospital with Covid-19, according to exposure to each class of antidepressants. The shaded areas represent pointwise 95% confidence intervals.

**Table 2.**
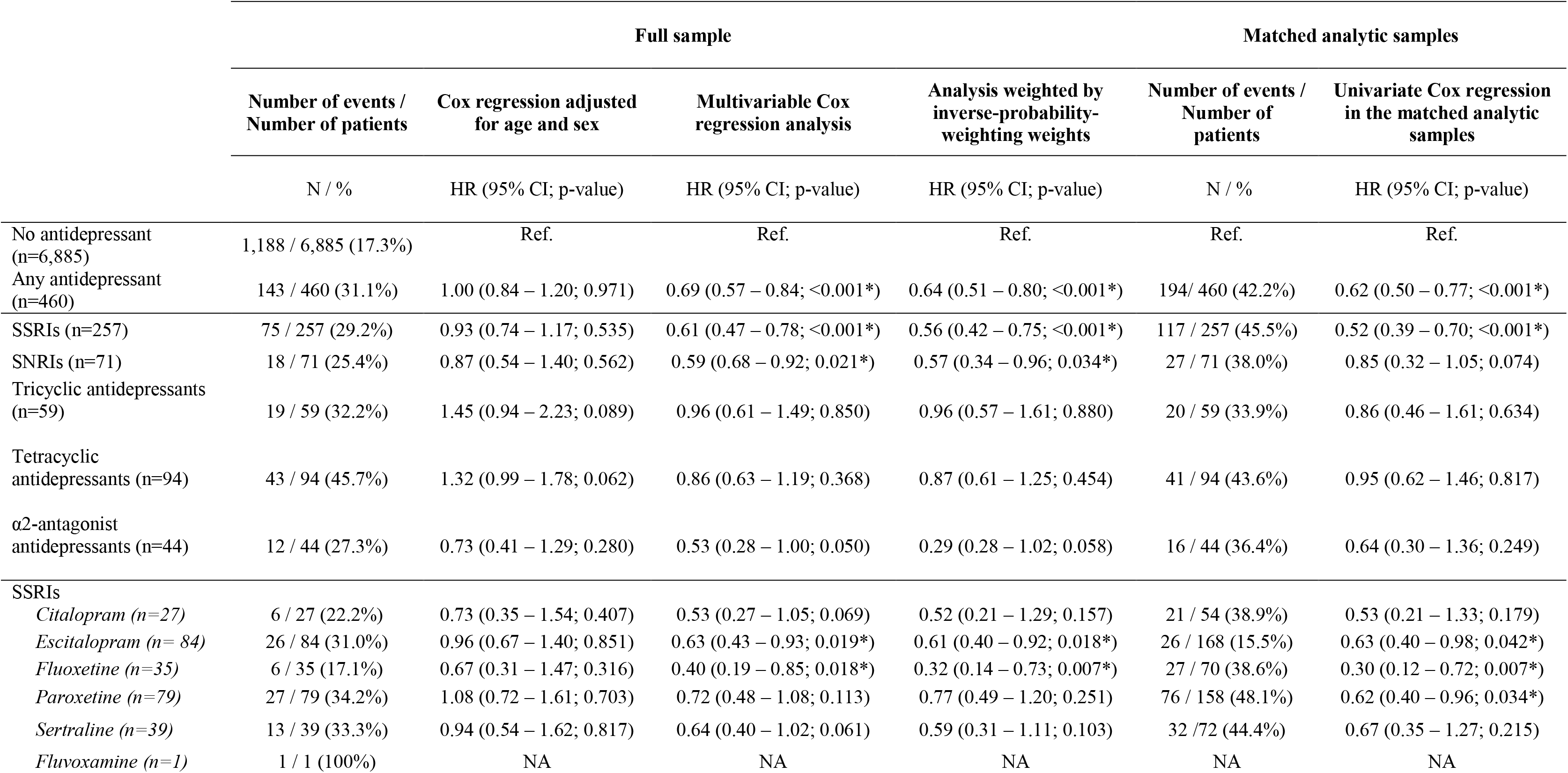
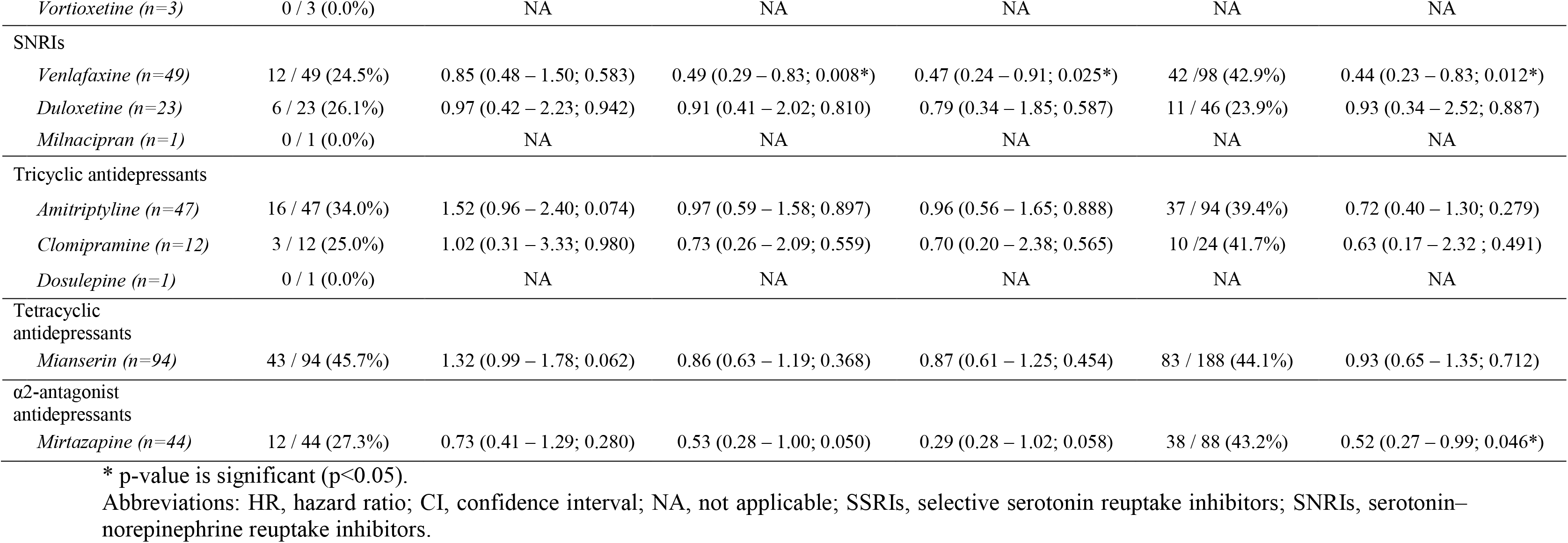
Associations of the use of any antidepressant, each antidepressant class and each individual treatment with the composite endpoint of intubation or death in the full sample and in the matched analytic samples.

|  | Full sample |  |  |  | Matched analytic samples |  |
| --- | --- | --- | --- | --- | --- | --- |
|  | Number of events /<br>Number of patients | Cox regression adjusted<br>for age and sex | Multivariable Cox<br>regression analysis | Analysis weighted by<br>inverse-probability-weighting weights | Number of events /<br>Number of<br>patients | Univariate Cox regression<br>in the matched analytic<br>samples |
|  | N / % | HR (95% CI; p-value) | HR (95% CI; p-value) | HR (95% CI; p-value) | N / % | HR (95% CI; p-value) |
| No antidepressant<br>(n=6,885) | 1,188 / 6,885 (17.3%) | Ref. | Ref. | Ref. | Ref. | Ref. |
| Any antidepressant<br>(n=460) | 143 / 460 (31.1%) | 1.00 (0.84 – 1.20; 0.971) | 0.69 (0.57 – 0.84; <0.001*) | 0.64 (0.51 – 0.80; <0.001*) | 194 / 460 (42.2%) | 0.62 (0.50 – 0.77; <0.001*) |
| SSRIs (n=257) | 75 / 257 (29.2%) | 0.93 (0.74 – 1.17; 0.535) | 0.61 (0.47 – 0.78; <0.001*) | 0.56 (0.42 – 0.75; <0.001*) | 117 / 257 (45.5%) | 0.52 (0.39 – 0.70; <0.001*) |
| SNRIs (n=71) | 18 / 71 (25.4%) | 0.87 (0.54 – 1.40; 0.562) | 0.59 (0.68 – 0.92; 0.021*) | 0.57 (0.34 – 0.96; 0.034*) | 27 / 71 (38.0%) | 0.85 (0.32 – 1.05; 0.074) |
| Tricyclic antidepressants<br>(n=59) | 19 / 59 (32.2%) | 1.45 (0.94 – 2.23; 0.089) | 0.96 (0.61 – 1.49; 0.850) | 0.96 (0.57 – 1.61; 0.880) | 20 / 59 (33.9%) | 0.86 (0.46 – 1.61; 0.634) |
| Tetracyclic<br>antidepressants (n=94) | 43 / 94 (45.7%) | 1.32 (0.99 – 1.78; 0.062) | 0.86 (0.63 – 1.19; 0.368) | 0.87 (0.61 – 1.25; 0.454) | 41 / 94 (43.6%) | 0.95 (0.62 – 1.46; 0.817) |
| α2-antagonist<br>antidepressants (n=44) | 12 / 44 (27.3%) | 0.73 (0.41 – 1.29; 0.280) | 0.53 (0.28 – 1.00; 0.050) | 0.29 (0.28 – 1.02; 0.058) | 16 / 44 (36.4%) | 0.64 (0.30 – 1.36; 0.249) |
| SSRIs |  |  |  |  |  |  |
| *Citalopram (n=27)* | 6 / 27 (22.2%) | 0.73 (0.35 – 1.54; 0.407) | 0.53 (0.27 – 1.05; 0.069) | 0.52 (0.21 – 1.29; 0.157) | 21 / 54 (38.9%) | 0.53 (0.21 – 1.33; 0.179) |
| *Escitalopram (n= 84)* | 26 / 84 (31.0%) | 0.96 (0.67 – 1.40; 0.851) | 0.63 (0.43 – 0.93; 0.019*) | 0.61 (0.40 – 0.92; 0.018*) | 26 / 168 (15.5%) | 0.63 (0.40 – 0.98; 0.042*) |
| *Fluoxetine (n=35)* | 6 / 35 (17.1%) | 0.67 (0.31 – 1.47; 0.316) | 0.40 (0.19 – 0.85; 0.018*) | 0.32 (0.14 – 0.73; 0.007*) | 27 / 70 (38.6%) | 0.30 (0.12 – 0.72; 0.007*) |
| *Paroxetine (n=79)* | 27 / 79 (34.2%) | 1.08 (0.72 – 1.61; 0.703) | 0.72 (0.48 – 1.08; 0.113) | 0.77 (0.49 – 1.20; 0.251) | 76 / 158 (48.1%) | 0.62 (0.40 – 0.96; 0.034*) |
| *Sertraline (n=39)* | 13 / 39 (33.3%) | 0.94 (0.54 – 1.62; 0.817) | 0.64 (0.40 – 1.02; 0.061) | 0.59 (0.31 – 1.11; 0.103) | 32 / 72 (44.4%) | 0.67 (0.35 – 1.27; 0.215) |
| *Fluvoxamine (n=1)* | 1 / 1 (100%) | NA | NA | NA | NA | NA |
| *Vortioxetine (n=3)* | 0 / 3 (0.0%) | NA | NA | NA | NA | NA |
| SNRIs |  |  |  |  |  |  |
| *Venlafaxine (n=49)* | 12 / 49 (24.5%) | 0.85 (0.48 – 1.50; 0.583) | 0.49 (0.29 – 0.83; 0.008*) | 0.47 (0.24 – 0.91; 0.025*) | 42 / 98 (42.9%) | 0.44 (0.23 – 0.83; 0.012*) |
| *Duloxetine (n=23)* | 6 / 23 (26.1%) | 0.97 (0.42 – 2.23; 0.942) | 0.91 (0.41 – 2.02; 0.810) | 0.79 (0.34 – 1.85; 0.587) | 11 / 46 (23.9%) | 0.93 (0.34 – 2.52; 0.887) |
| *Milnacipran (n=1)* | 0 / 1 (0.0%) | NA | NA | NA | NA | NA |
| Tricyclic antidepressants |  |  |  |  |  |  |
| *Amitriptyline (n=47)* | 16 / 47 (34.0%) | 1.52 (0.96 – 2.40; 0.074) | 0.97 (0.59 – 1.58; 0.897) | 0.96 (0.56 – 1.65; 0.888) | 37 / 94 (39.4%) | 0.72 (0.40 – 1.30; 0.279) |
| *Clomipramine (n=12)* | 3 / 12 (25.0%) | 1.02 (0.31 – 3.33; 0.980) | 0.73 (0.26 – 2.09; 0.559) | 0.70 (0.20 – 2.38; 0.565) | 10 / 24 (41.7%) | 0.63 (0.17 – 2.32 ; 0.491) |
| *Dosulepine (n=1)* | 0 / 1 (0.0%) | NA | NA | NA | NA | NA |
| Tetracyclic antidepressants |  |  |  |  |  |  |
| *Mianserin (n=94)* | 43 / 94 (45.7%) | 1.32 (0.99 – 1.78; 0.062) | 0.86 (0.63 – 1.19; 0.368) | 0.87 (0.61 – 1.25; 0.454) | 83 / 188 (44.1%) | 0.93 (0.65 – 1.35; 0.712) |
| α2-antagonist antidepressants | | | | | | |
| *Mirtazapine (n=44)* | 12 / 44 (27.3%) | 0.73 (0.41 – 1.29; 0.280) | 0.53 (0.28 – 1.00; 0.050) | 0.29 (0.28 – 1.02; 0.058) | 38 / 88 (43.2%) | 0.52 (0.27 – 0.99; 0.046*) |

\* p-value is significant (p<0.05).
Abbreviations: HR, hazard ratio; CI, confidence interval; NA, not applicable; SSRIs, selective serotonin reuptake inhibitors; SNRIs, serotonin–norepinephrine reuptake inhibitors.

In sensitivity analyses, multivariable Cox regression models in the full sample yielded similar results, as did univariate Cox regression models in the matched analytic samples, except for the association between SNRI use and the primary endpoint, which was not significant (HR, 0.85; 95% CI, 0.32 to 1.05, p=0.074), and exposures to paroxetine (HR, 0.62; 95% CI, 0.40 to 0.96, p=0.034) and mirtazapine (HR, 0.52; 95% CI, 0.27 to 0.99, p=0.046), which were significantly associated with reduced risk of intubation or death in these analyses (**Table 2; eFigures 1 to 5**).

Exposure to higher rather than lower doses of antidepressants was not significantly associated with risk of intubation or death, except for tetracyclic antidepressants, for which higher doses were associated with an increased risk (**eTable 15**). Exposure to a combination of antidepressants was not significantly associated with this risk compared to exposure to only one antidepressant (**eTable 16**).

When using intubation and death as separate endpoints, we found that exposures to any antidepressant and any SSRI, but not to other classes of antidepressants, were significantly associated with lower risks (**eTable 17**).

Among the 593 patients admitted to ICUs, antidepressant use was significantly associated with reduced risk of intubation or death in the primary multivariable Cox model with inverse probability weighting, but not in other multivariable analyses, possibly because of a lack of statistical power, as only 28 (4.7%) of these patients received an antidepressant (**eTable 18**).

Finally, among patients with COVID-19 not exposed to any antidepressant during the hospitalization, there was no significant association between antidepressant use or SSRI use in the 3 months before hospital admission and the primary endpoint (**eTable 19**).

## Discussion

In this multicenter observational study involving a large sample of patients admitted to the hospital with COVID-19, we found that antidepressant use, and specifically exposures to SSRIs and to the SSRIs fluoxetine and escitalopram and the SNRI venlafaxine, were significantly and substantially associated with reduced risk of intubation or death. The association between SSRI use and this risk was observed at a mean dosage of 21.4 (SD=13.6) fluoxetine-equivalent milligrams. These findings should be interpreted with caution due to the observational design and the fact that this is, to our knowledge, the first study examining the efficacy of antidepressants in a clinical population of patients with COVID-19. However, our results suggest that SSRI use may be associated with reduced risk of intubation or death in hospitalized patients with COVID-19.

In the analyses, we tried to minimize the effects of confounding in several different ways. First, we used multivariable regression models with inverse probability weighting to minimize the effects of confounding by indication.^21^ ^22^ We also performed sensitivity analyses, including multivariable Cox regression models and univariate Cox regression models in matched analytic samples, that showed similar results. Second, although some amount of unmeasured confounding may remain, our analyses adjusted for numerous potential confounders, including sex, age, obesity, smoking status, any medical condition associated with increased risk of severe COVID-19, any medication prescribed according to compassionate use or as part of a clinical trial, markers of clinical and biological severity of COVID-19, mood or anxiety or other current psychiatric disorder, and any prescribed benzodiazepine or Z-drug, mood stabilizer, and antipsychotic medication. Furthermore, our findings indicate that these associations were not significantly different across multiple subgroups defined by baseline characteristics, except for biological severity of COVID-19 at hospital admission, and only observed for patients receiving these medications during the visit. Third, these associations remained significant for any antidepressant use and SSRI use when using death and intubation as separate endpoints. Finally, the low number of patients with antidepressants admitted to ICUs (N=28) in our study and the significant association between antidepressant use and reduced risk of intubation or death among patients admitted to ICUs in the propensity score analysis with inverse probability weighting further give strength to our conclusion.

Additional limitations of our study include missing data for some baseline characteristic variables, including baseline clinical and biological severity of COVID-19, which may be explained by the overwhelming of all hospital units during the COVID-19 peak incidence, and potential for inaccuracies in the electronic health records in this context, such as the possible lack of documentation of illnesses or medications, or the misidentification of treatments’ mode of administration (e.g., dosage, frequency), especially for hand-written medical prescriptions. Furthermore, type I error inflation due to multiple testing may have occurred in our study. However, our analyses were exploratory, and results were similar across different statistical approaches. Finally, despite the multicenter design, our results may not be generalizable to other settings or regions.

In this multicenter observational retrospective study involving patients admitted to the hospital with COVID-19, SSRI use at usual antidepressant doses during the visit was associated with lower risk of intubation or death. Double-blind controlled randomized clinical trials of these medications for COVID-19 are needed.

## Data Availability

Data from the AP-HP Health Data Warehouse can be obtained at https://eds.aphp.fr//.

## Acknowledgments

The authors thank the EDS APHP Covid consortium integrating the APHP Health Data Warehouse team as well as all the APHP staff and volunteers who contributed to the implementation of the EDS-Covid database and operating solutions for this database.

Collaborators of the EDS APHP Covid consortium are: Pierre-Yves ANCEL, Alain BAUCHET, Nathanaël BEEKER, Vincent BENOIT, Mélodie BERNAUX, Ali BELLAMINE, Romain BEY, Aurélie BOURMAUD, Stéphane BREANT, Anita BURGUN, Fabrice CARRAT, Charlotte CAUCHETEUX, Julien CHAMP, Sylvie CORMONT, Christel DANIEL, Julien DUBIEL, Catherine DUCLOAS, Loic ESTEVE, Marie FRANK, Nicolas GARCELON, Alexandre GRAMFORT, Nicolas GRIFFON, Olivier GRISEL, Martin GUILBAUD, Claire HASSEN-KHODJA, François HEMERY, Martin HILKA, Anne Sophie JANNOT, Jerome LAMBERT, Richard LAYESE, Judith LEBLANC, Léo LEBOUTER, Guillaume LEMAITRE, Damien LEPROVOST, Ivan LERNER, Kankoe LEVI SALLAH, Aurélien MAIRE, Marie-France MAMZER, Patricia MARTEL, Arthur MENSCH, Thomas MOREAU, Antoine NEURAZ, Nina ORLOVA, Nicolas PARIS, Bastien RANCE, Hélène RAVERA, Antoine ROZES, Elisa SALAMANCA, Arnaud SANDRIN, Patricia SERRE, Xavier TANNIER, Jean-Marc TRELUYER, Damien VAN GYSEL, Gaël VAROQUAUX, Jill Jen VIE, Maxime WACK, Perceval WAJSBURT, Demian WASSERMANN, Eric ZAPLETAL.

## Contributors

NH designed the study, performed statistical analyses, and wrote the first draft of the manuscript. MSR contributed to study design, performed statistical analyses and critically revised the manuscript. FL contributed to study design and critically revised the manuscript for scientific content. RV contributed to statistical analyses and critically revised the manuscript for scientific content. NB and ASJ contributed to study design and critically revised the manuscript for scientific content. NB, ASJ, AN, NP, CD, AG, GL, MB, and AB contributed to database build process. AN, CB, MO, CL, GA, NP, CD, AG, GL, MB, and AB critically revised the manuscript for scientific content. NH is the guarantor. The corresponding author attests that all listed authors meet authorship criteria and that no others meeting the criteria have been omitted.

## Ethical approval

This observational study using routinely collected data received approval from the Institutional Review Board of the AP-HP clinical data warehouse (decision CSE-20-20_COVID19, IRB00011591). AP-HP clinical Data Warehouse initiative ensures patients’ information and informed consent regarding the different approved studies through a transparency portal in accordance with European Regulation on data protection and authorization n°1980120 from National Commission for Information Technology and Civil Liberties (CNIL). All procedures related to this work adhered to the ethical standards of the relevant national and institutional committees on human experimentation and with the Helsinki Declaration of 1975, as revised in 2008.

## Data sharing

Data from the AP-HP Health Data Warehouse can be obtained at https://eds.aphp.fr//.

<u>The corresponding author affirms that the manuscript is an honest, accurate, and transparent account of the study being reported. No important aspects of the study have been omitted and any discrepancies from the study as planned have been disclosed.</u>

## Dissemination to participants and related patient and public communities

No patients were involved in setting the research question or the outcome measures, nor were they involved in the design and implementation of the study. We plan to disseminate these findings to participants through the AP-HP website and to the general public in a press release.

## Exclusive licence

The Corresponding Author has the right to grant on behalf of all authors and does grant on behalf of all authors, an exclusive licence on a worldwide basis to the BMJ Publishing Group Ltd to permit this article (if accepted) to be published in BMJ editions and any other BMJPGL products and sublicences such use and exploit all subsidiary rights, as set out in our licence.

---

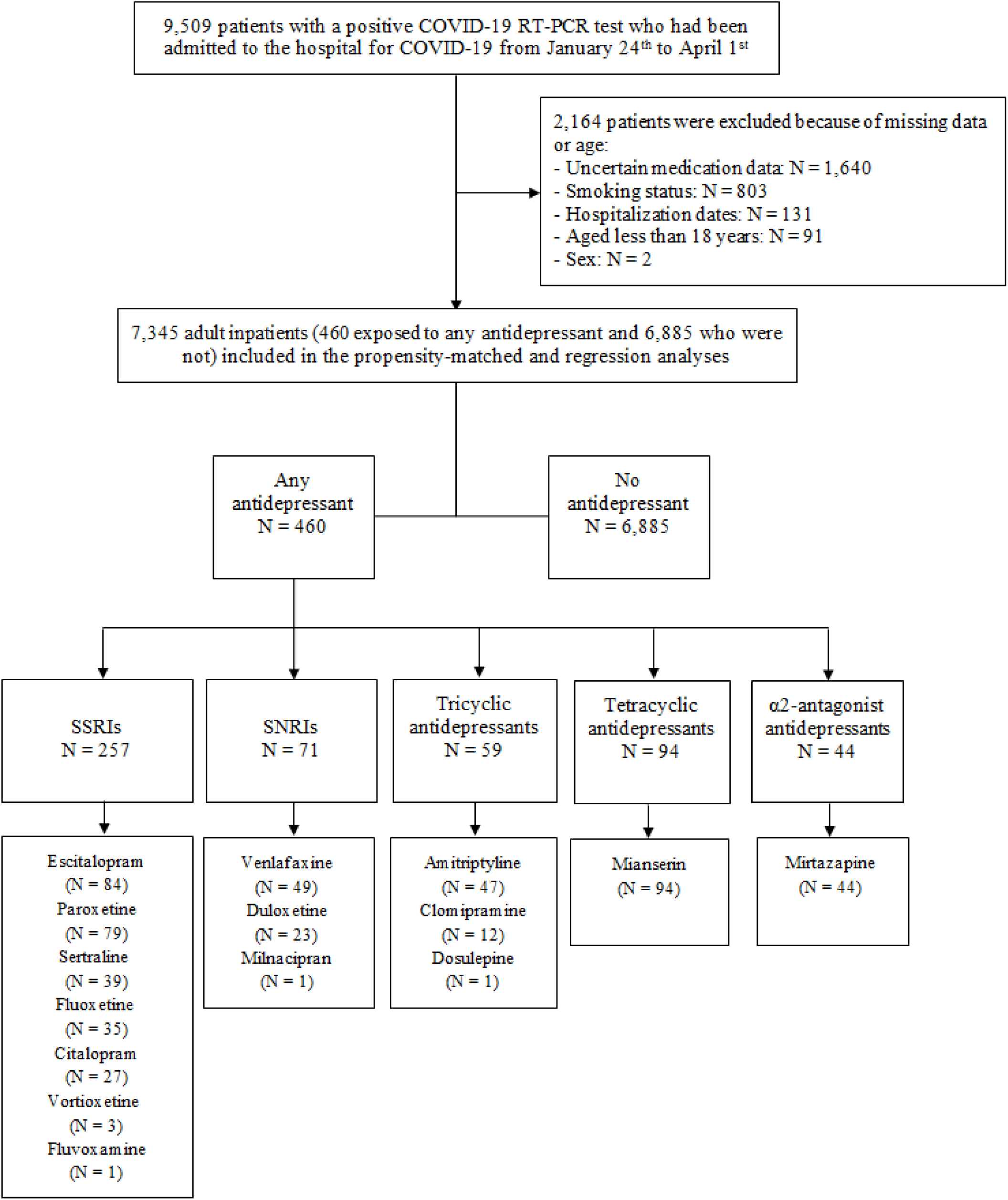
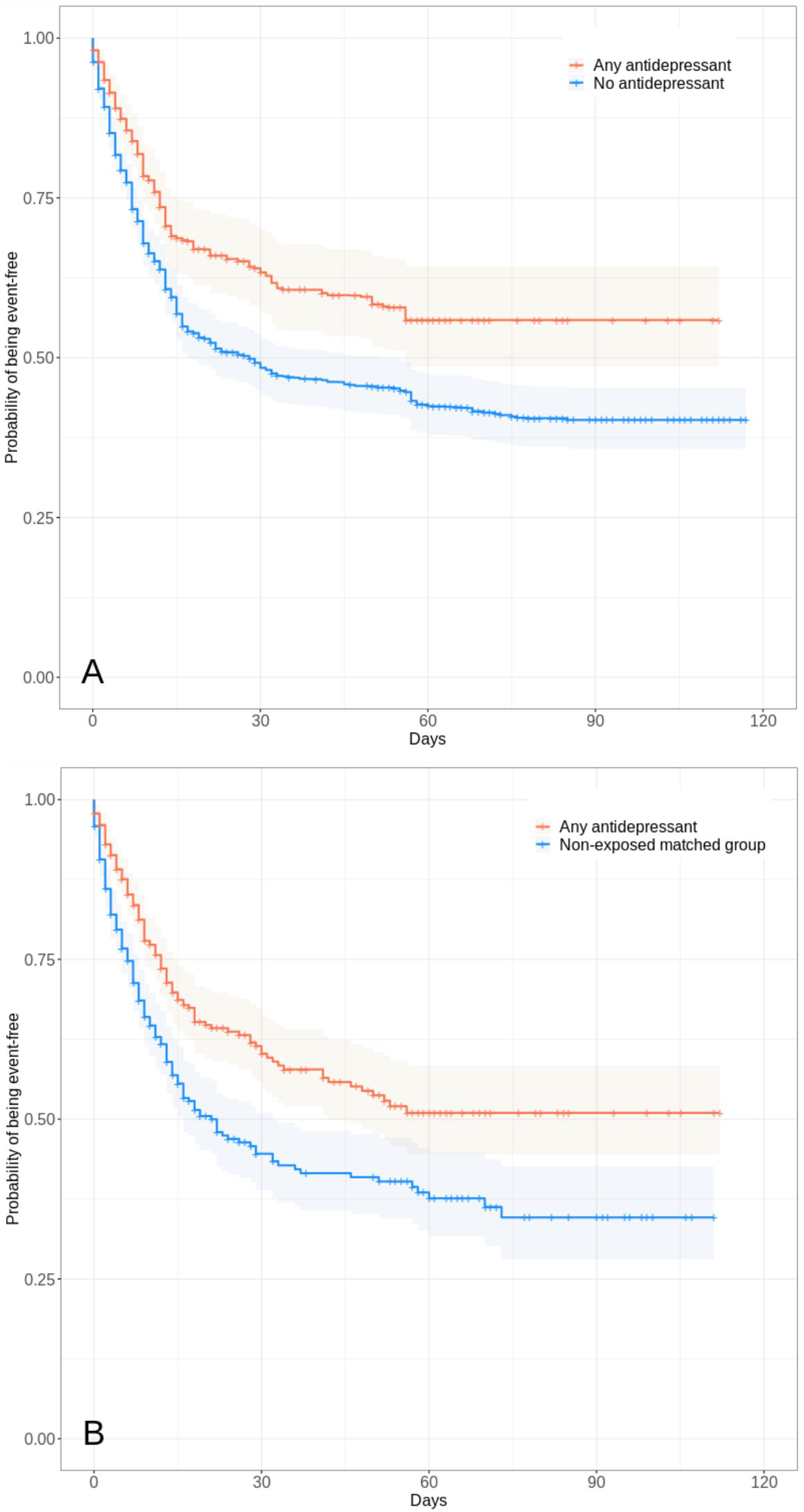
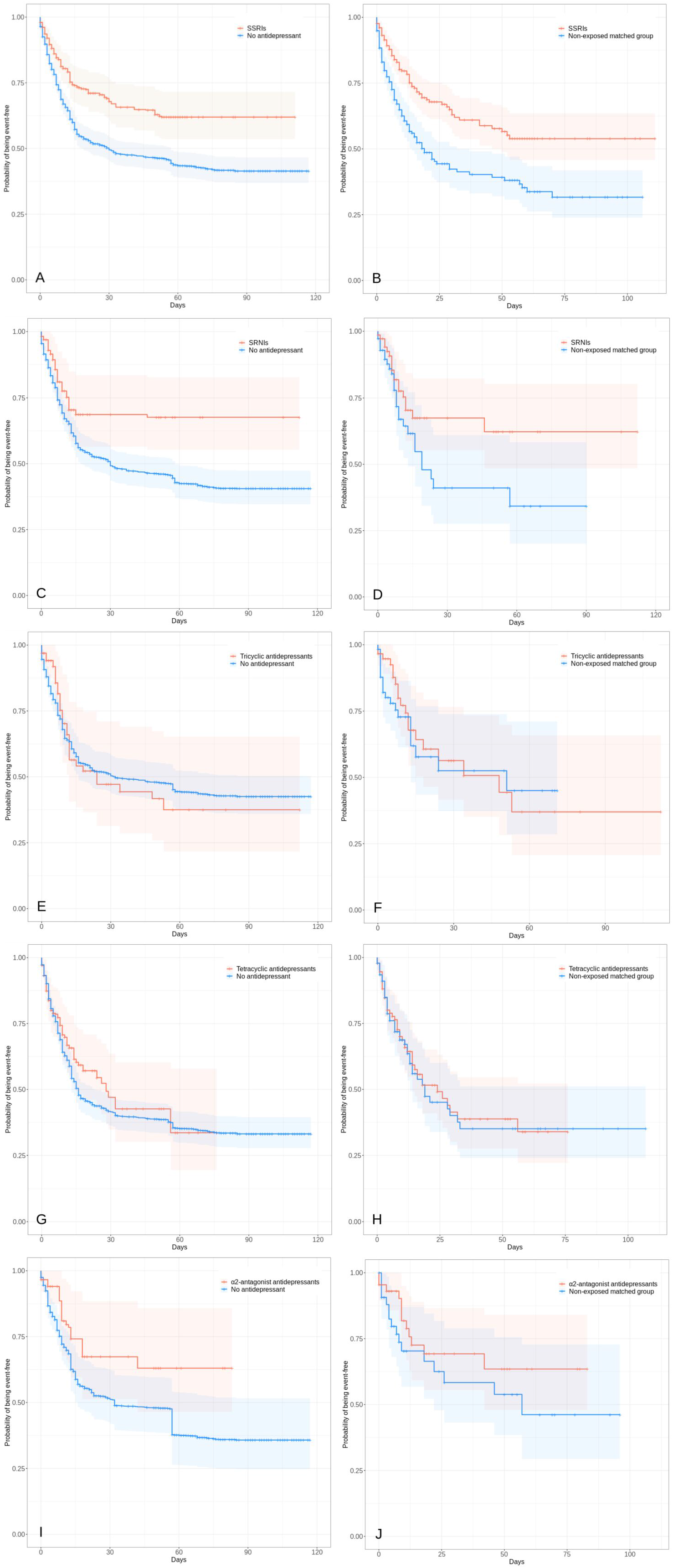
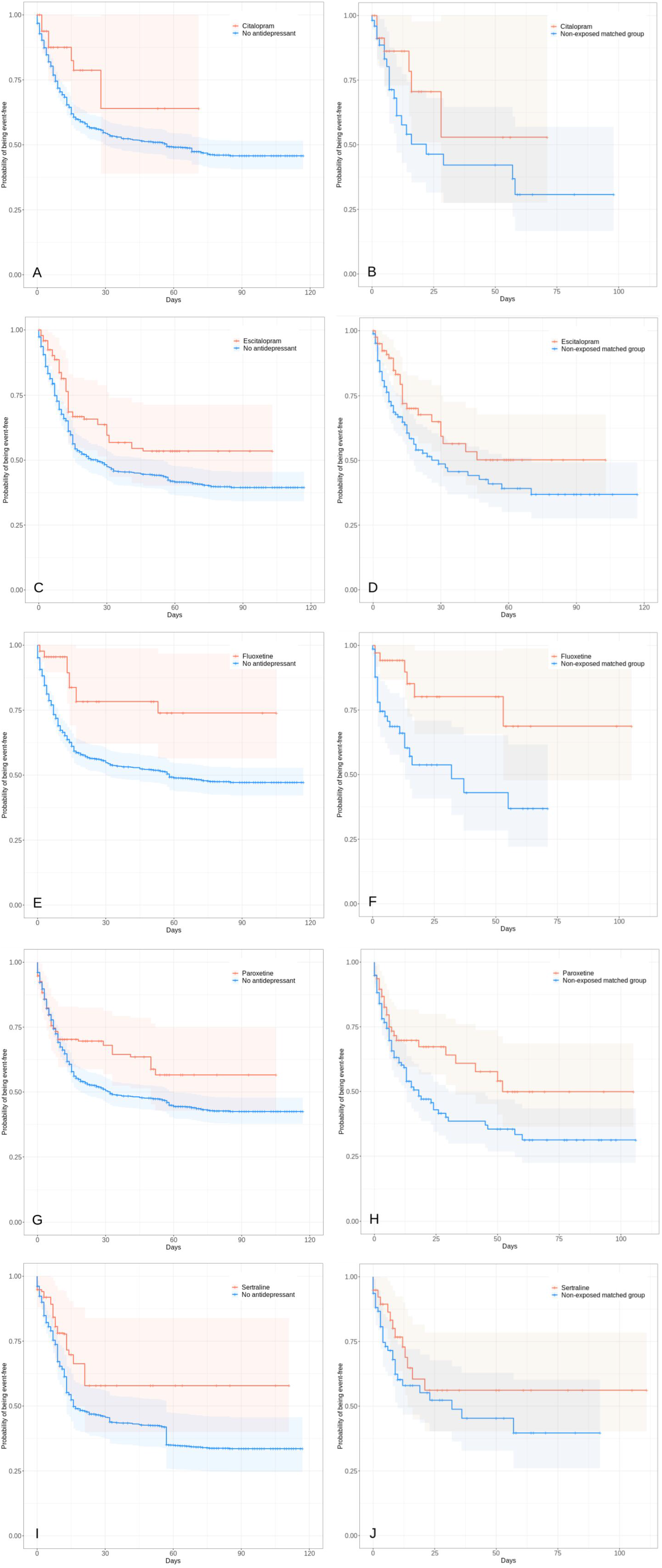
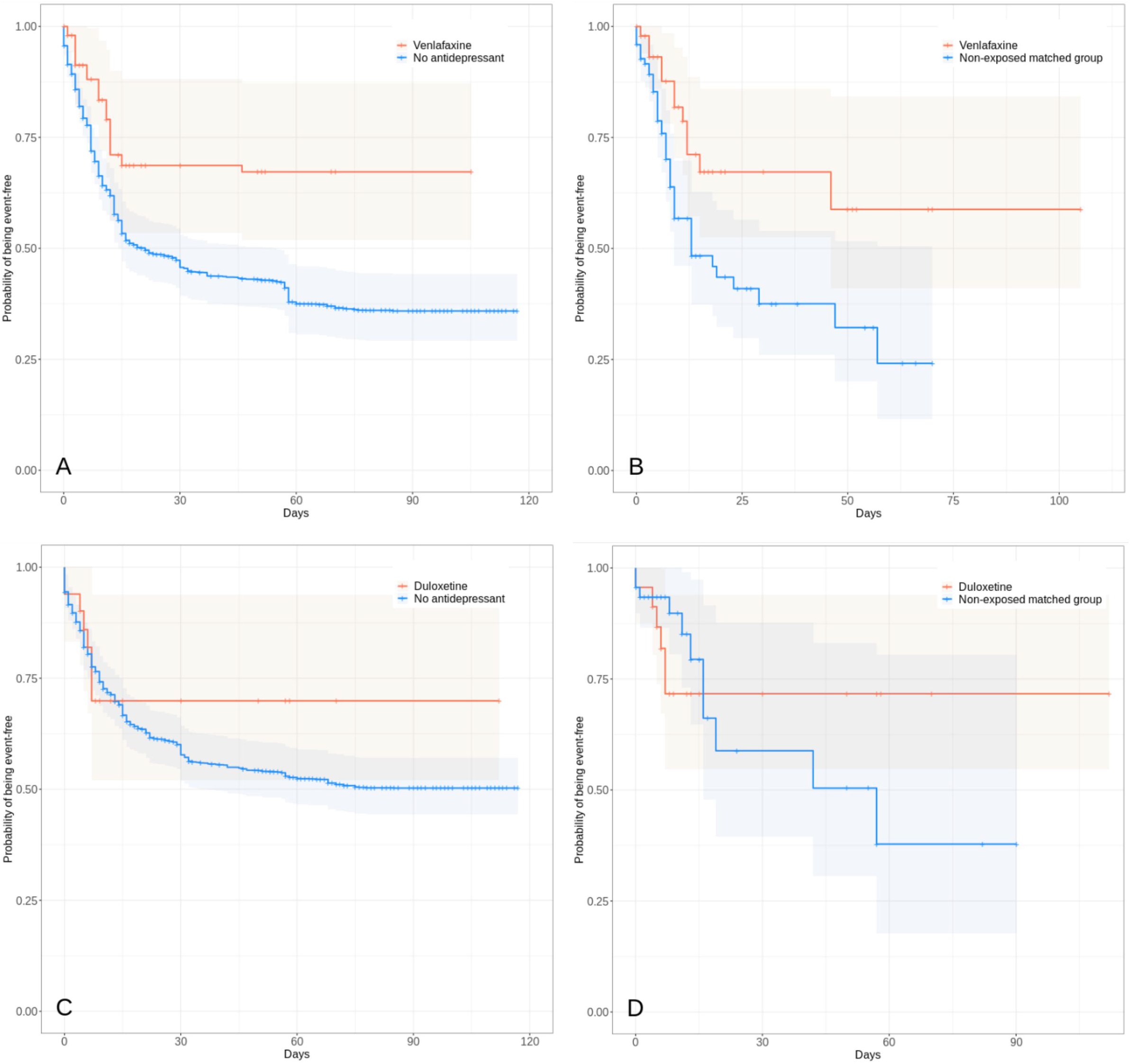
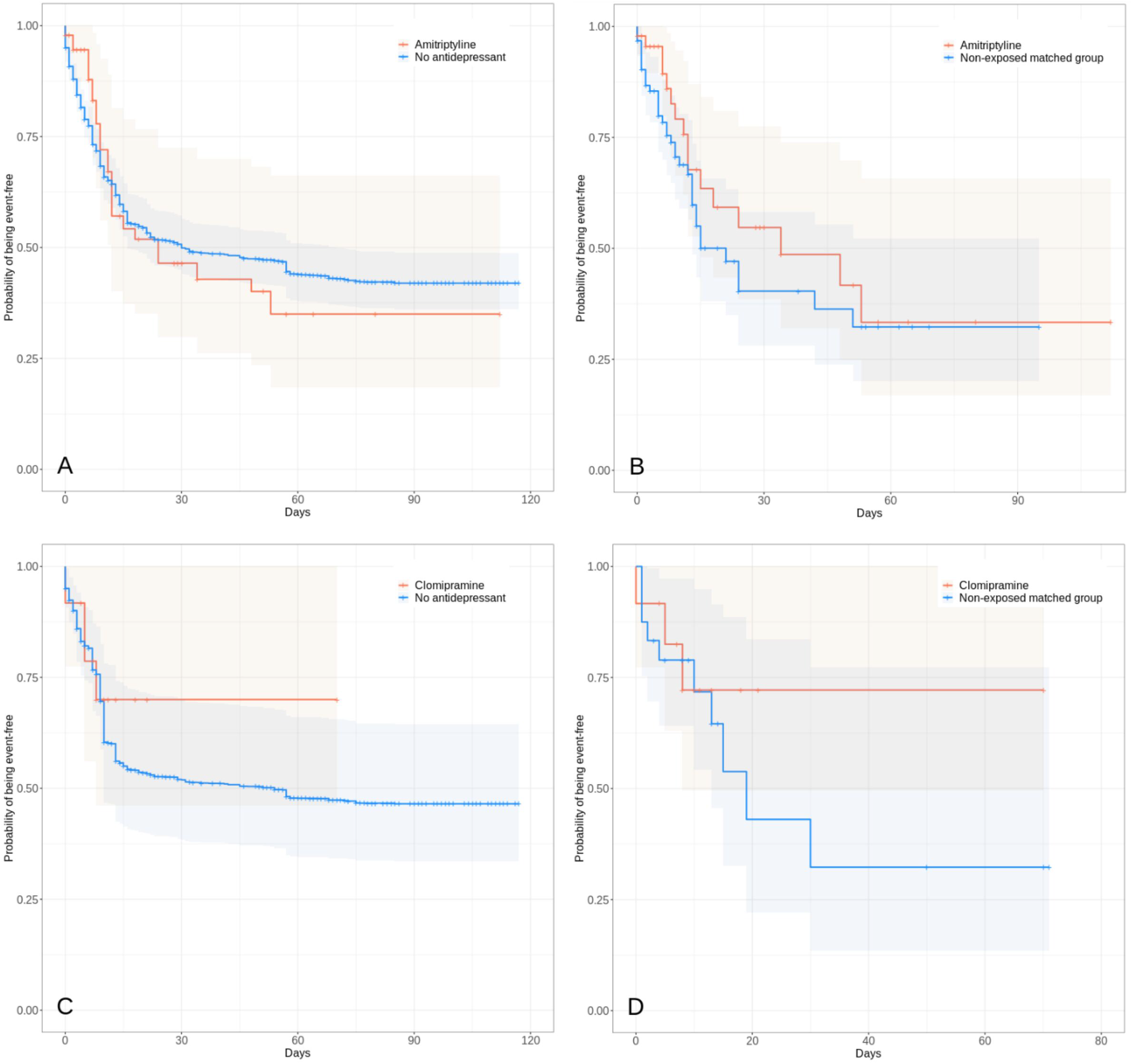
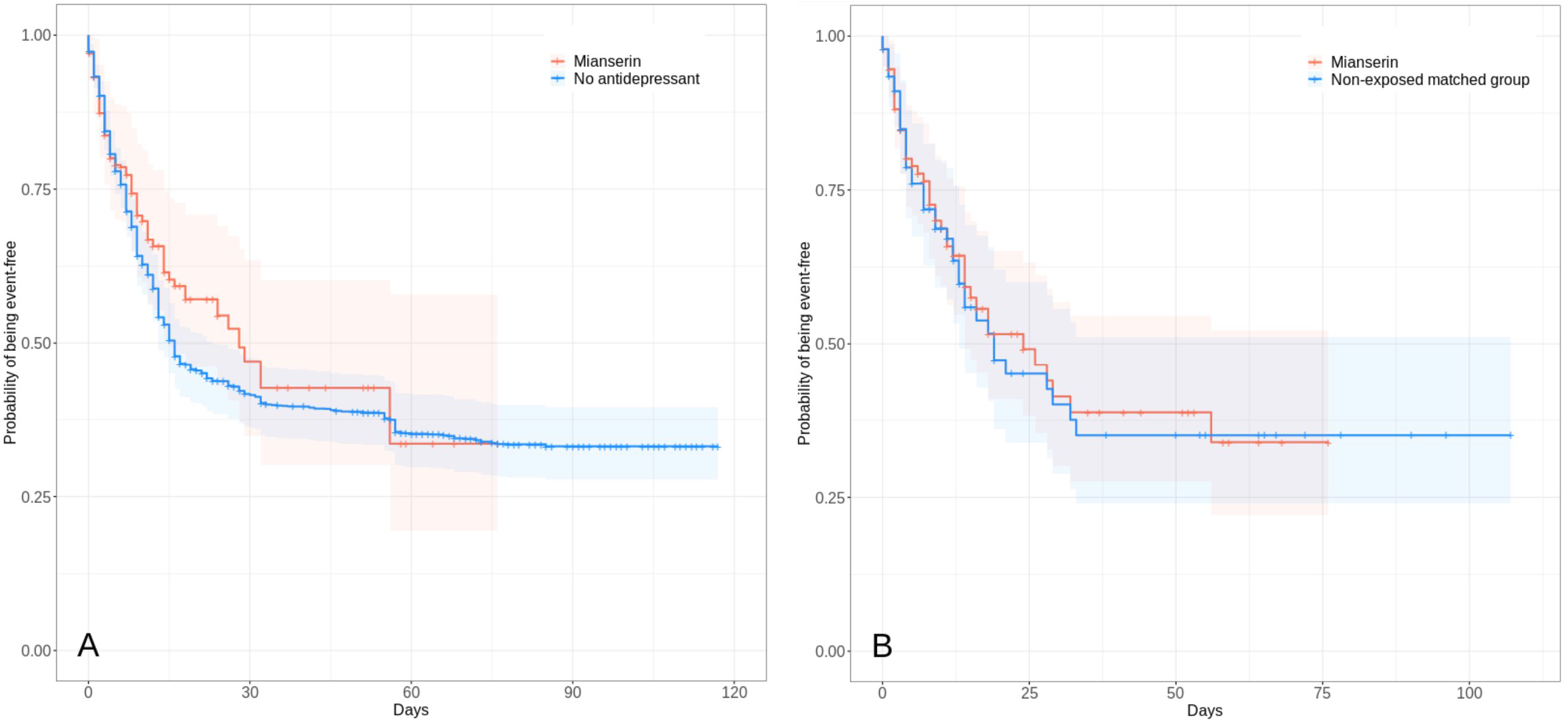
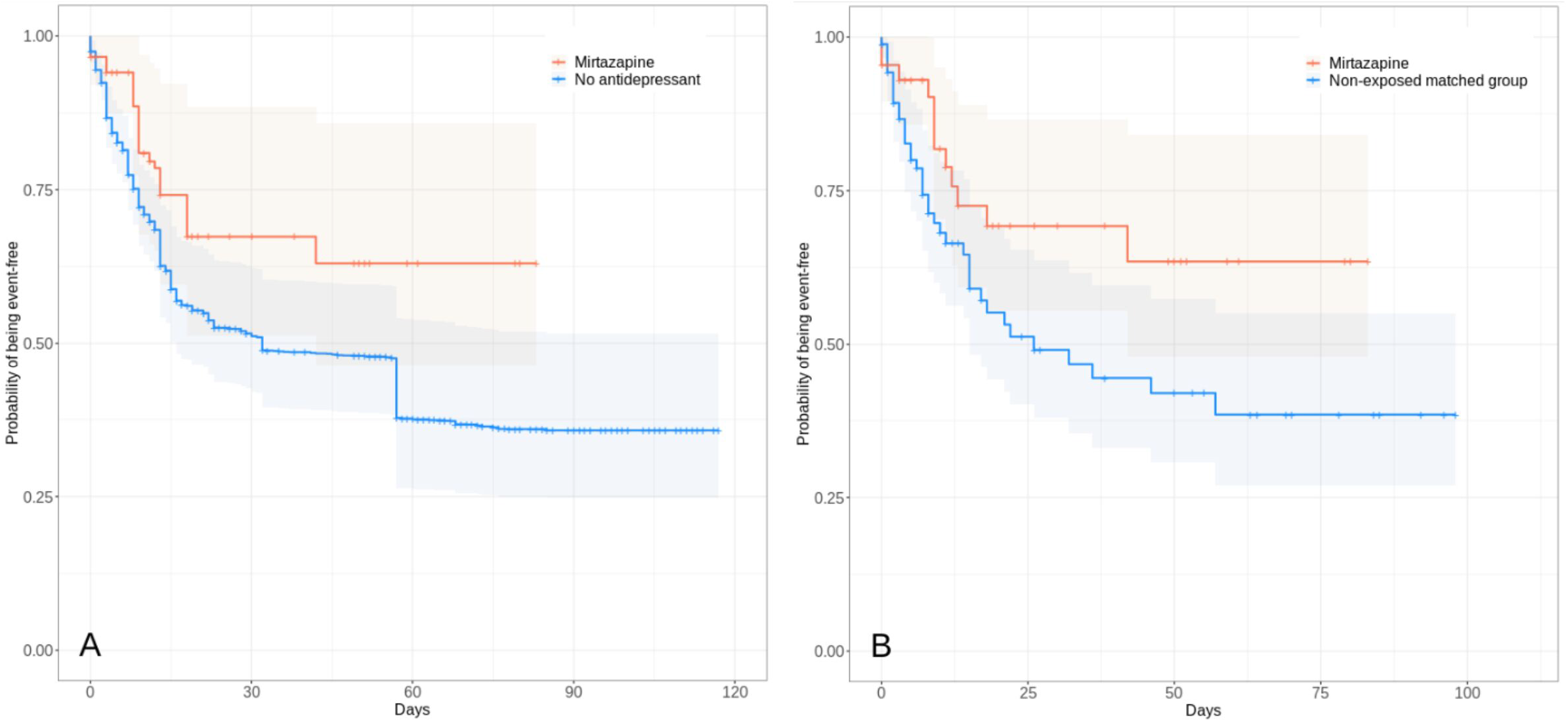

